# Analysis of the time evolution of SARS-CoV-2 lethality rate in Italy: Evidence of an unaltered virus potency

**DOI:** 10.1101/2020.06.12.20129387

**Authors:** Nicole Balasco, Vincenzo d’Alessandro, Giovanni Smaldone, Luigi Vitagliano

## Abstract

In recent months, the entire world is facing a dramatic health emergency caused by the diffusion of a hitherto unknown coronavirus (SARS-CoV-2). Despite the efforts, the understanding of the many facets of the pandemic is still rather limited. In the present manuscript, we have monitored the evolution of the lethality rate in Italy by using the data collected over the last three months. Our data indicate that there is a striking correlation between the number of infected people of a certain week and the deaths of the following one. Despite the overall simplicity of the applied approach and its many approximations, the analysis of the Italian scenario provides some interesting insights into the pandemic. Indeed, we have found that the lethality rate is virtually unchanged over the last two months. This implies that the reduction of the deaths is strictly connected to the decrease of cases. Unfortunately, the present study does not support the idea that the virus potency has lowered in the last weeks, as our data demonstrate that the likelihood of a fatal outcome after the infection has not decreased in the recent outbreak evolution. Moreover, we show that the lethality rate is still very high in the country (≈13.5%). Since this number is remarkably higher if compared to the actual lethality estimates made worldwide, this finding suggests that the number of detected cases may be a gross underestimation of the actual infected people, likely due to the presence of a significant number of non-symptomatic or paucisymptomatic individuals in the population.

## Introduction

In China, the last part of 2019 has been characterized by the detection of a novel severe acute respiratory syndrome of unknown origin that did not respond to common treatments of respiratory illnesses. The causative agent of this rapidly-diffusing disease has been identified in an unknown coronavirus that has been denoted as SARS-CoV-2 (Brussow 2020 and references therein, Chen et al. 2020). The unravelling of the virus genome has highlighted that SARS-CoV-2 shares remarkable similarities with the coronaviruses responsible for SARS and MERS syndromes, but also some clear distinctive features.

According to the data reported by Worldometer^*a*^, the spread of the virus throughout the world has caused a pandemic that currently involves more than 200 countries of five continents. SARS-CoV-2 has hitherto infected over seven million people worldwide with the death of more than 400k individuals. Despite the huge efforts made, preventive and broadly effective therapeutic approaches to combat the virus and to cure the related respiratory syndrome are currently not available. It is a common opinion that this situation will not change in the coming months. To limit the pandemic, social distance restrictions and, frequently, lockdowns of large regions and countries have been imposed at almost planetary scale. Although this strategy, where applied, has significantly mitigated the virus diffusion, its economic and social costs are becoming progressively unsustainable for many communities. Therefore, there is an urgent but largely unmet need to gain a deep understanding of the many facets of this global and dramatic health emergency.

A striking complication of such a new and unforeseen scenario is that the enormous collection of related data, poorly validated by the scientific community and rapidly diffused by the media, has frequently produced disputes and controversies. Moreover, it is virtually impossible for scientists working in this field to keep updated with the huge amount of literature produced. This sort of ‘live science’ with a weakly-filtered osmosis of information from the lab bench to public has often generated confusion that could eventually lead to a general distrust for science.

In the SARS-CoV-2 pandemic, Italy has a prominent position since it has been the locus of the first major outbreak among Western countries (Ding and Gao 2020, Di Lorenzo and Di Trolio 2020, Giangreco 2020, Iosa 2020, Livingston and Bucher 2020, Michelozzi et al. 2020, Onder et al. 2020, Rovetta et al. 2020). Moreover, the Lethality Rate (i.e., number of deaths over number of infected people), also referred to as Case Fatality Rate or CFR (Modi et al. 2020, De Natale et al. 2020), of the infection in Italy was significantly higher than that detected in most of other regions/countries. To face the emergency, the Italian authorities activated a severe lockdown over the entire country (March 9^th^) that was gradually released two months later. This initiative had a remarkable impact on the virus diffusion, although the effects emerged rather slowly. The country is presumably experiencing the end of the pandemic, as indicated by the reduced number of cases and deaths. However, it is important to note that the number of infected people (cases) and fatalities is far from being negligible and any kind of intervention should be carefully evaluated. In this scenario, there is a strong national debate about the actions, even if any, which have to be taken to cope with the infection. In the last weeks, one of the most controversial issues is related to the possible loss of potency of the virus that could have been caused, among others, by factors such as the changed weather conditions and/or possible SARS-CoV-2 mutations recently emerged from the virus variants^b^. In the framework of our research activities focused on the classification and analysis of aminoacid mutations, we here evaluate whether a variation of virus potency could be detected at epidemiologic level by using very simple approaches. In particular, we report an extremely simple analysis of the temporal link between the number of SARS-CoV-2 cases and the number of deceased people in Italy. Despite the considerable simplicity of the method, we derive interesting connections between different events in the infection evolution. Unfortunately, our analyses do not corroborate the idea that the virus has recently lost potency during its spread in the Italian population.

## Methodology

Cases, deaths, and tests related to SARS-CoV-2 were daily collected from the bulletin of the Italian Civil Protection^*c*^ (Protezione Civile) and manually curated for *a posteriori* correction (**Table S1**). The values are virtually identical to those reported by Worldometers^*d*^. All collected data (cases, deaths, and cases/tests) were organized and averaged in a week-based grouping. Starting from February 24^th^ we collected data over 14 weeks from W_0_ (Feb 24^th^ – Mar 1^st^) to W_13_ (May 25^th^ – May 31^st^). The full list of the week definition is reported in **Table S2**. Since data became more complete and reliable only after some days from the beginning of the pandemic, most of the statistics were done starting from W_1_ (Mar 2^nd^ – Mar 8^th^). The grouped and averaged daily data in a week-based manner is reported for each ensemble (cases, deaths, and tests) in **Table S3**.

We here define a Weekly Lethality Rate (WLR), i.e., a weekly CFR, as the ratio between the weekly average number of deaths and the weekly average number of cases. In our analyses, to establish a causative link between these two parameters, the numbers of cases and deaths in the ratio are not necessarily corresponding to the same week. Most of the analyses were performed by assuming a one-week shift (1WS) between deaths and cases. As consequence, the ratio (R_i_) was calculated dividing the deaths of a given week (W_i_) by the cases of the preceding week (W_i-1_); accordingly, R_2_, R_3_, … and R_13_ represent the deaths/cases ratios given by W_2_/W_1_, W_3_/W_2_ …, and W_13_/W_12_, respectively. We occasionally used different schemes to calculate the ratio either with no shift (deaths and cases of the same week - 0WS) or with a shift of two weeks (2WS). The values of ratios of all the schemes are reported in **Table S4**.

## Results and Discussion

### Overall trends of cases and deaths

The number of cases, deaths, and tests were collected from the daily reports of the Protezione Civile and reported in **Table S1** and **Figure 1** (see the Methodology section for further details). The analysis of the time evolution of cases (**Figure 1A**) reveals that the peak was reached in the time interval March 20^th^-28^th^. The curve is indicative of a rapid progression of cases, followed, after the peak, by a rather slow reduction of infected people that is still ongoing. No major variation of the trend is observed as the consequence of the release of the lockdown (May 18^th^), although a final assessment on this issue should be made in a longer time scale. The inspection of the curve of the deceased people indicates that it is endowed with similar features. A rapidly growing phase is indeed followed by a smooth decrease of daily deaths (**Figure 1B**). The peak of the curve is observed in the March 26^th^ - April 1^st^ time span.

**Figure 1.**
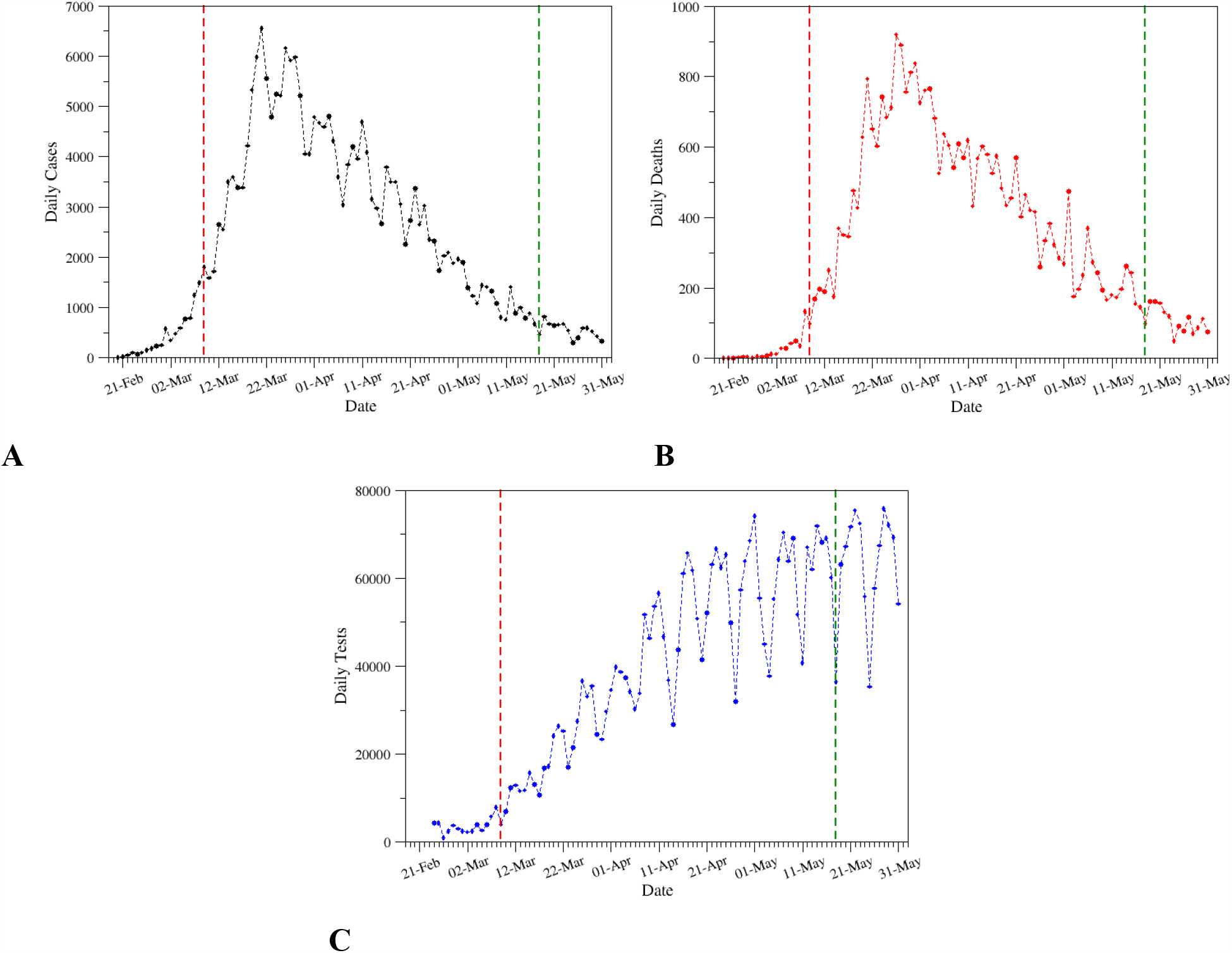
Daily cases (A), deaths (B), and tests (C) in the interval Feb 20^th^ - May 31^st^, as retrieved from the bulletin of the Protezione Civile. The red and green vertical dashed lines correspond to the dates of the lockdown in Italy (March 9^th^) and its release (May 18^th^), respectively. The data used to generate the graphs are reported in **Table S1**.

### Identification of a quantitative correlation between the trends

The close similarity between the curves of cases and deaths prompted us to look for a quantitative temporal correlations between these events. A normalization of the two datasets was preliminarily performed by dividing all points for the maximum value of each ensemble (**Table S3**). The inspection of **Figure 2A** that reports these normalized values highlights the analogies of the two curves and suggests that they are shifted one to each other. As expected, the curve of deaths is delayed compared to that of cases.

**Figure 2.**
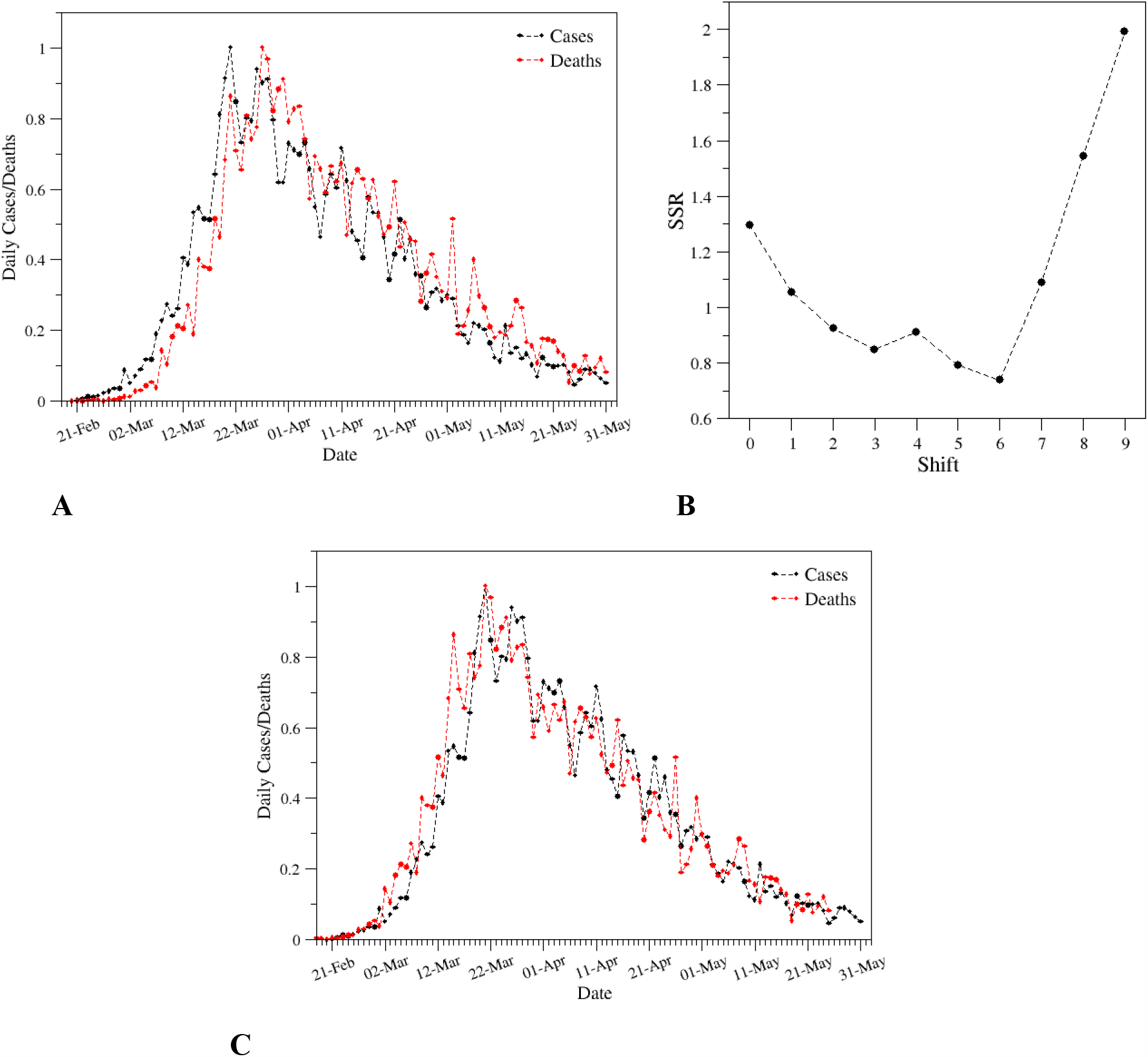
Comparison of the evolution of the number of cases (black) and deaths (red) upon normalization of the curves (A). The normalization was performed by dividing the actual values for the maximum of each ensemble. The sum of squared residuals (SSR) is reported in the panel B as a function of the number of day shift applied to the curve of deaths. The optimal overlay of the curves (shift = 6) is shown in panel C.

To quantitatively compare the observed trends, we systematically shifted the curve of deaths and computed the sum of squared residuals (SSR) of the overlaid curves. As indicated by **Figure 2B** that reports the SSR values as function of the shifted days, the optimal fitting of the data is achieved by applying a six-day shift to the curve of deaths. This value, which represents the average delay between the swab outcome and the corresponding death, is compatible with the average shift of eleven days between the insurgence of the symptoms and the fatal outcome reported by the Italian National Health Institute - ISS (Report of May 28^th^)^e^.

Since all data exhibit a clear weekday dependence (**Figure 1**), we grouped them in a weekly-based manner. In particular, we preliminarily computed the total week number of cases/deaths by summing up the daily data. Then, average daily values were obtained dividing these numbers by seven. For each ensemble, we normalized the values by dividing all data by the maximum value. As shown in **Figure 3**, the resulting curves display better defined trends compared to the daily ones as they exhibit a monotonic behavior before and after the peak.

**Figure 3.**
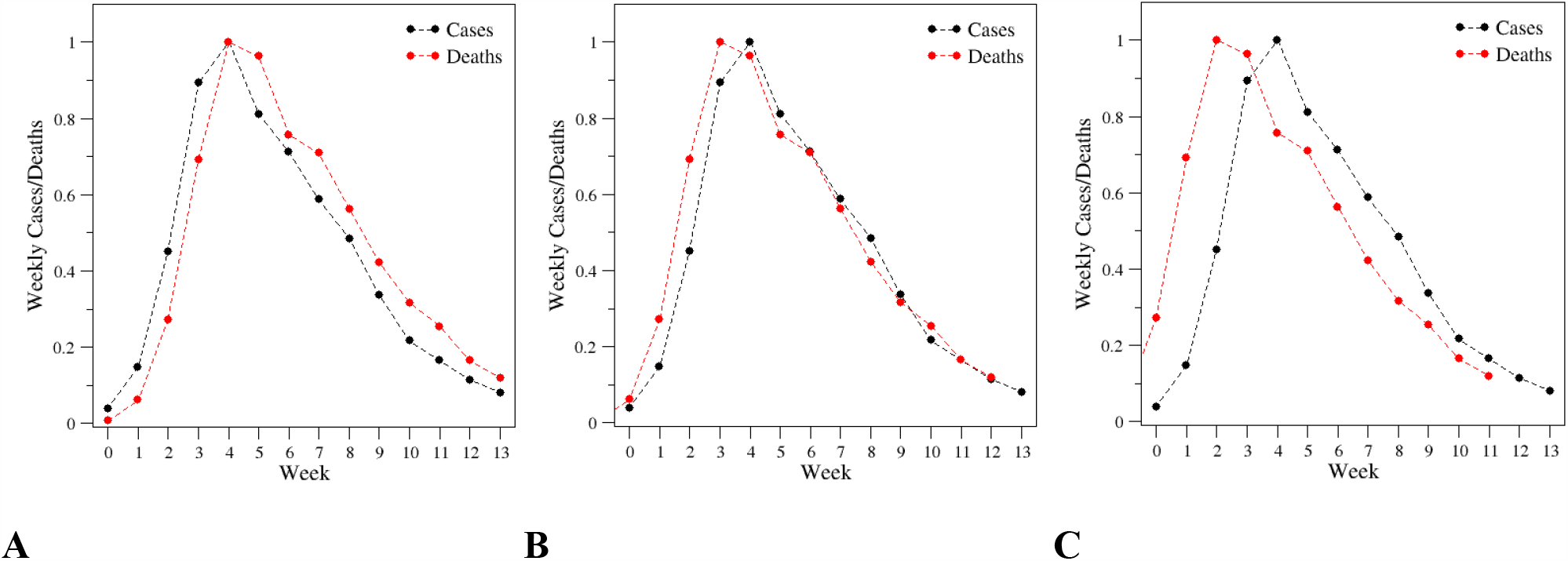
Evolution of weekly averaged cases (black) and deaths (red) upon normalization of the data (A). The normalization was performed by dividing the actual values by the maximum of each ensemble. In panel B and C, the overlay obtained after the shift of one or two week(s), respectively, is reported. The data illustrated in panel A are listed in **Table S3**.

As expected, the curve of deaths presents a temporal delay when compared to that of the cases. By adopting the approach used for the daily data, we shifted the curve of the deaths by a fixed number of weeks. In line with the results illustrated above, the comparative analysis of **Figures 3B** and **3C** indicates that the optimal fitting is obtained by a shift of one week (1WS). It is important to note that the fitting obtained with a single-week shift leads to a remarkably good overlap of the curves in the region that follows the peaks. The fitting is slightly worse in the initial weeks. This is likely due to the poor estimate of the cases in this period ascribable to the extremely low number of tests performed (**Figure 1C**).

Using these weekly-organized data, we also monitored the ratio of the number of cases (or deaths) between consecutive weeks (**Figure 4** and **Table S5**). The evolution of this ratio is rather similar in the two ensembles of data. In both cases, it is characterized by a rapid drop in the initial weeks of the infection followed by fairly constant values in more recent weeks. This behavior is the consequence of the almost linear decrease of cases and deaths in the recent phase of the pandemic (**Figures 1** and **3**).

**Figure 4.**
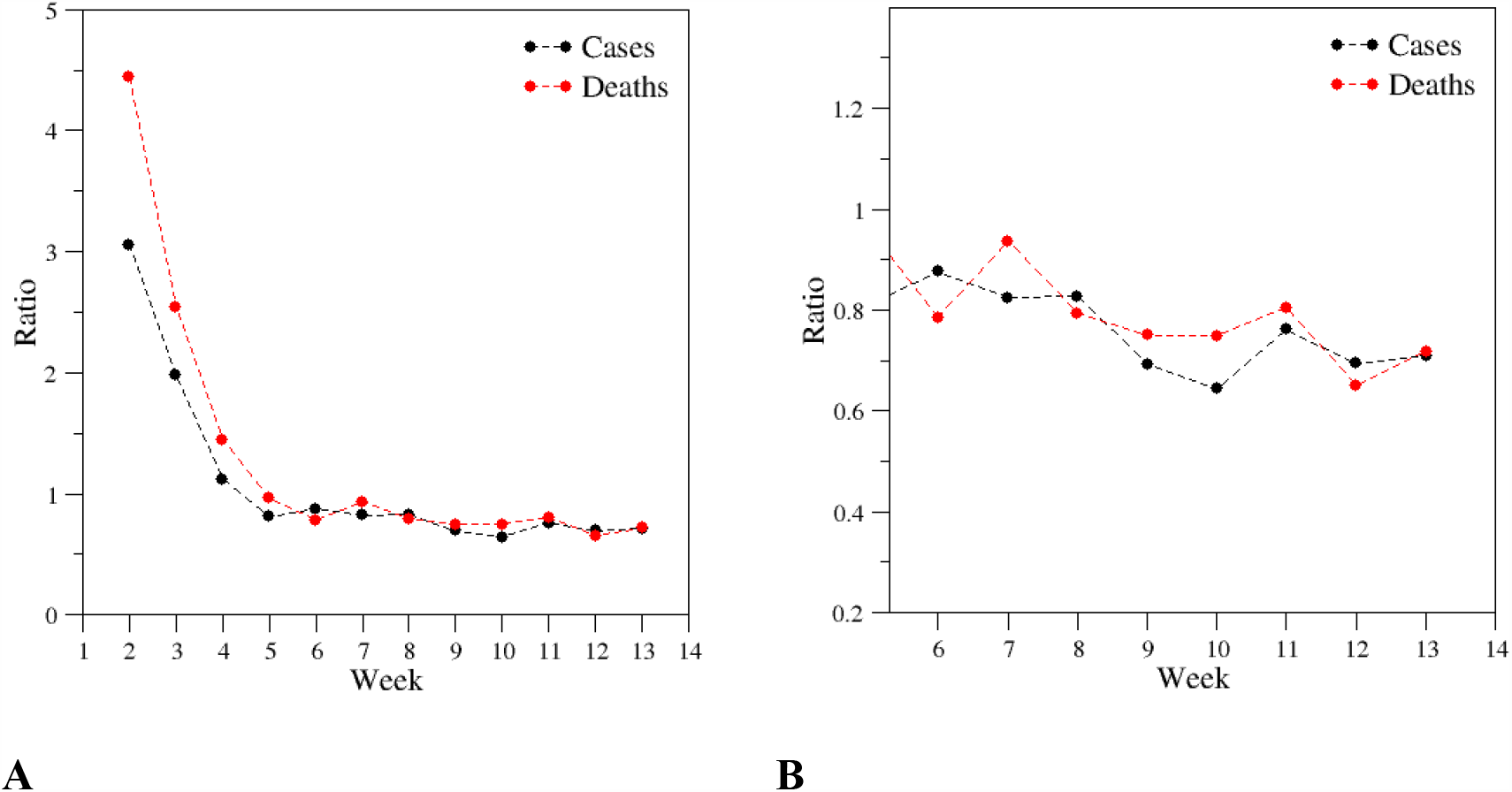
Ratio of cases (black line) and deaths (red line) between consecutive weeks. Values were computed by dividing the cases (or deaths) of a given week (W_i_) by the cases (or deaths) of one week (W_i-1_) before. For example, the ratio #2 is calculated by dividing the cases (or deaths) of W_2_ by those of W_1_. The panel B is a snapshot of panel A aimed at highlighting the trends of the last weeks. The data used to generate the graphs are reported in **Table S5**.

### Time evolution of the lethality

To gain further insights into the SARS-CoV-2 diffusion and lethality in Italy, we exploited the week-based data illustrated in the previous paragraph to calculate the WLR. In particular, we calculated the WLR values as the ratios between weekly deaths/cases assuming a one week shift (1WS) scheme, i.e., the deaths are counted in the week following the week used to estimate the cases. The analysis of **Figure 5** reporting the time evolution of the WLR indicates a reduction of the ratio in the initial weeks of the infection that, starting from the week five, is followed by a fairly constant trend (black line). The rather large WLR values detected in the first weeks (range 15 - 25%) are likely due to a substantial underestimation of the cases in that phase of the infection. Indeed, as shown in **Figure 5A**, in the initial weeks a large portion of the tests were positive (red line). As the consequence of more extensive testing campaigns, the cases/tests ratio has dropped from 25.8% (the maximum value) to 0.7% (last week - W_13_).

**Figure 5.**
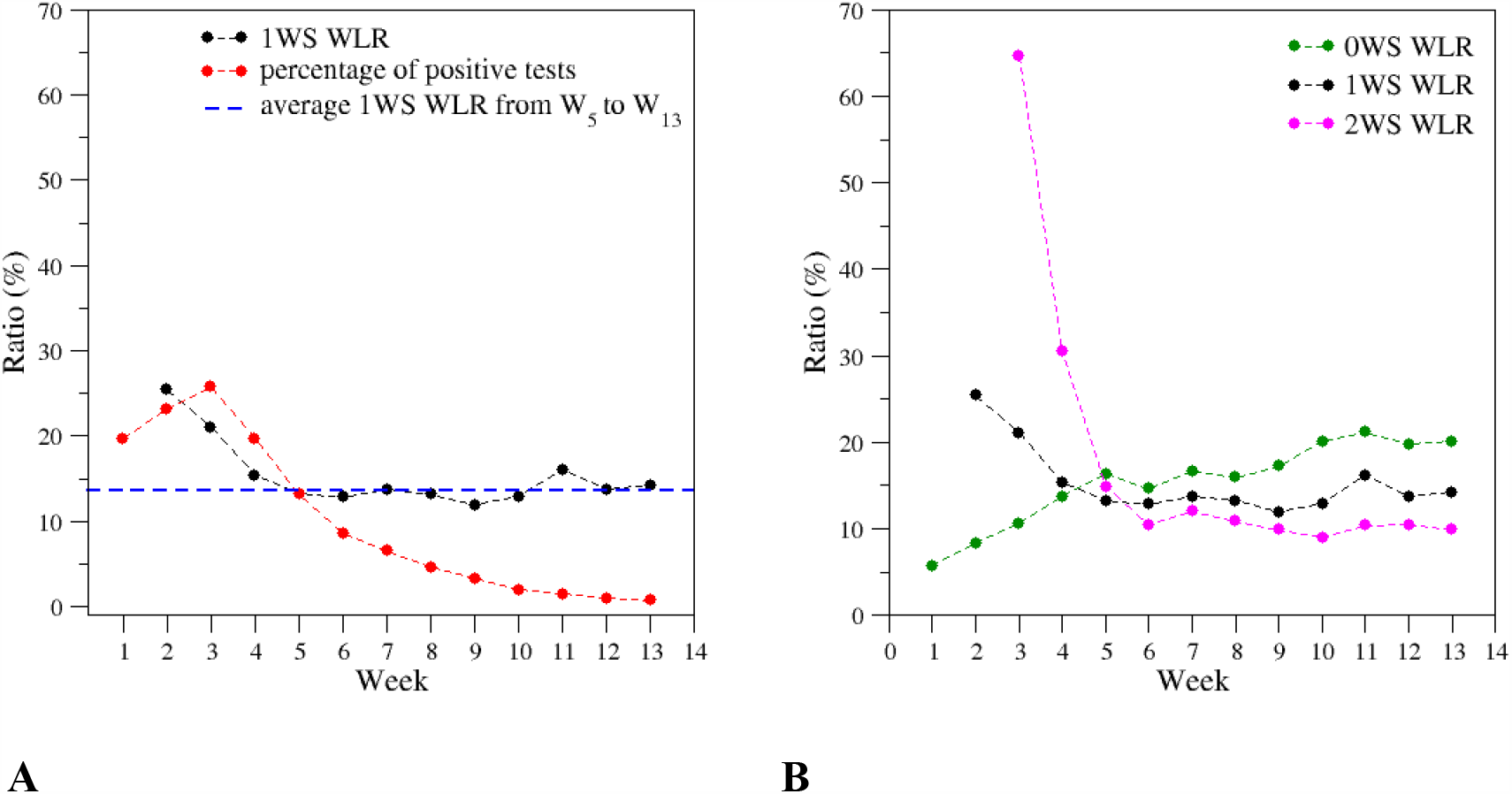
Time evolution of the Weekly Lethality Rate (WLR) based on the assumption of a one-week shift (1WS) (black line). The panel also reports the percentage of positive tests over the total number of tests (red line). The dashed blue line represents a horizontal line with an intercept of 13.6. This is the average 1WS WLR from W_5_ to W_13_. In the panel B, a comparison of the WLR based on the 1WS (black) scheme with the 0WS (green) and 2WS (magenta) ones is reported. The data used to generate the graphs are reported in **Table S4**.

Taking into account the constancy of WLR from week 5 to 13 that entirely cover April and May, we averaged the corresponding values. The resulting mean value of this ensemble is 13.6% with a standard deviation of 1.2%. A close inspection of **Figure 5A**, in which this average value is reported as a horizontal line, indicates a slight increase of the WLR in the last three weeks. Indeed, the average values (and standard deviations) calculated considering weeks from W_5_ to W_10_ or from W_11_ to W_13_ are 13.0 (± 0.6)% or 14.7 (± 1.2)%, respectively. However, it should be noted that the very low number of ratios considered does not warrant a significance to this slight increase. Additional data should be collected to validate or confute this observation.

The observed constancy of the WLR values after the initial decrease is somehow surprising as the percentage of positive tests is continuously decreasing. The reduction of the positive tests is also observed if a single test per individual is considered (**Figure S1** and **Table S4**).

Finally, we also analyzed the trends of the WLR values by assuming alternative schemes for the shift. In particular, we calculated new ratios by not considering any shift between deaths and cases (0WS) or by assuming a two weeks shift (2WS). As shown in **Figure 5B**, the trends emerged for 1WS are clearly observed also for 0WS and 2WS. Indeed, a reduction of the WLR values is observed in the initial phase of the infection that is followed by a *plateau*. Obviously, the mean value of WLR in the constant region obtained applying these schemes is different. Indeed, since after the peak the number of deaths decreases, the longer is the shift the smaller is the ratio.

### Validation of the constant model

To quantitatively check and validate the robustness of the model of the WLR evolution obtained with the present analysis, we predicted its value for W_12_ and W_13_ starting from the average WLR detected for the ensemble W_5_-W_11_, 13.4 (± 1.3)%. Using this value, we estimated the number of deaths of W_12_ and W_13_ by considering the cases of W_11_ and W_12_, respectively. The predicted number of deaths of W_12_ is 853 (6365×0.134). The actual deaths of this week are 877. The relative error of this prediction is 2.7%. The same approach used for W13 predicts 592 deaths (4423×0.134) with a relative error of 5.9% considering the actual 630 fatalities of this week. The very low error associated with these predictions witnesses the reliability of the illustrated approach. It is worth mentioning that in both cases we underestimate the deaths. This is not surprising if we consider that the WLR values are slightly increasing in the very last weeks of the infection.

## Concluding remarks

In the present manuscript, we monitored the evolution of the fatality rate in Italy by using the data collected over the last three months. Our data indicate that there is a striking correlation between the cases of one week and the deaths in the following one. Despite the overall simplicity of our approach and its many approximations, as for example the statistical assumption of a fixed time between deaths and cases, the analysis of the Italian scenario provides some interesting insights into this dramatic health emergency. The first finding is that the Weekly Lethality Rate is virtually unchanged, at least over the last two months. Our analysis indicates that the reduction of the deaths is strictly connected to the decrease of cases produced by the restrictions imposed by the National and local authorities. Unfortunately, the present analysis does not support the idea that the virus potency has lowered in the last weeks as emerged from the media^*f*^, despite the detected tendency of the virus to mutate (van Dorp et al. 2020). Our data indicate that the likelihood of a fatal outcome after the infection has not decreased in the last weeks. Obviously, this analysis does not exclude that the variation of external factors, including weather conditions, has (or will have) an impact on the virus infectivity potential (contagiousness).

The average fatality rate we detected in the last weeks indicates that the SARS-CoV-2 lethality rate is still very high in the country (≈13.5%). This number is remarkably higher if compared to the actual lethality estimates made worldwide^*g*^ (Modi et al. 2020). If we assume that there have not been special conditions in Italy that favored such an unfortunate situation, this finding suggests that the number of detected cases is still a gross underestimation of the actual infected people due to the presence in the population of a significant number of non-symptomatic or paucisymptomatic individuals. A better estimate of the spread of the virus in the country could be obtained by focusing the attention on individuals that, in principle, are not expected to be positive. This will require extensive testing of people that do not display symptoms and did not have contacts with infected people.

## Data Availability

Data used for the analyses are reported in the supplementary material and are available from the authors

## Acknowledgments

The present activities have been performed in the framework of the project “RicErCa e sviluppO VERsus COVID19 in Campania RECOVER-COVID19” funded by the Campania Region.

## Footnotes

a From https://www.worldometers.info/

b From https://uk.reuters.com/article/uk-health-coronavirus-italy-virus/new-coronavirus-losing-potency-top-italian-doctor-says-idUKKBN2370OP; https://www.bloomberg.com/news/articles/2020-05-31/italy-s-new-coronavirus-cases-on-declining-trend-on-sunday; https://www.tvnz.co.nz/one-news/new-zealand/top-nz-immunologist-backs-theory-covid-19-losing-potency-could-become-common-cold-virus.

c From http://www.protezionecivile.gov.it/media-comunicazione

d From https://www.worldometers.info/coronavirus/italy

e From https://www.epicentro.iss.it/en/coronavirus/bollettino/Report-COVID-2019_28_May_2020.pdf

f From https://uk.reuters.com/article/uk-health-coronavirus-italy-virus/new-coronavirus-losing-potency-top-italian-doctor-says-idUKKBN2370OP; https://www.bloomberg.com/news/articles/2020-05-31/italy-s-new-coronavirus-cases-on-declining-trend-on-sunday; https://www.tvnz.co.nz/one-news/new-zealand/top-nz-immunologist-backs-theory-covid-19-losing-potency-could-become-common-cold-virus.

g From https://www.worldometers.info/coronavirus/coronavirus-death-rate/

## References

Brussow H. The Novel Coronavirus – Latest Findings. Microb. Biotechnol. 2020; 13, 819–828.

Chen Y, Liu Q, Guo D. Emerging Coronaviruses: Genome Structure, Replication, and Pathogenesis. J. Med. Virol. 2020 Apr;92(4):418–423.

De Natale G, Ricciardi V, De Luca G, De Natale D, Di Meglio G, Ferragamo A, Marchitelli V, Piccolo A, Scala A, Somma R, Spina E, Troise C. The COVID-19 Infection in Italy: A Statistical Study of an Abnormally Severe Disease. J. Clin. Med. 2020; 9(5), 1564.

Di Lorenzo G, Di Trolio R. Coronavirus Disease (COVID-19) in Italy: Analysis of Risk Factors and Proposed Remedial Measures. Front. Med. (Lausanne) 2020; 7: 140.

Ding Y, Gao L. An Evaluation of COVID-19 in Italy: A data-driven modeling analysis. 2020; doi: 10.21203/rs.3.rs-28146/v1

Giangreco G. Case fatality rate analysis of Italian COVID‐19 outbreak. J. Med. Virol. 2020; Apr 27: 10.1002/jmv.25894.

Iosa M, Paolucci S, Morone G. Covid-19: A Dynamic Analysis of Fatality Risk in Italy. Front. Med. (Lausanne) 2020; 7: 185.

Livingston E, Bucher K. Coronavirus disease 2019 (COVID-19) in Italy. JAMA. 2020;323(14):1335.

Michelozzi P, de’Donato F, Scortichini M, De Sario M, Noccioli F, Rossi P, Davoli M. Mortality impacts of the coronavirus disease (COVID-19) outbreak by sex and age: rapid mortality surveillance system, Italy, 1 February to 18 April 2020. Euro Surveill. 2020;25(19):pii=2000620.

Modi C, Boehm V, Ferraro S, Stein G, Seljak U. How deadly is COVID-19? A rigorous analysis of excess mortality and age-dependent fatality rates in Italy. MedRxiv. 2020; doi: https://doi.org/10.1101/2020.04.15.20067074.

Onder G, Rezza G, MD; Brusaferro S. Case-Fatality Rate and Characteristics of Patients Dying in Relation to COVID-19 in Italy. JAMA. 2020; 323(18):1775–1776.

Rovetta A, Bhagavathula AS, Castaldo L. Modelling the epidemiological trend and behavior of COVID-19 in Italy. MedRxiv. 2020. doi: https://doi.org/10.1101/2020.03.19.20038968.

van Dorp L, Acman M, Richard D, Shawd LP, Ford CE, Ormond L, Owen CJ, Panga J, Tan CCS, Boshiere FAT, Torres Ortiz A, Balloux F. Emergence of genomic diversity and recurrent mutations in SARS-CoV-2. Infect. Genet. Evol. 2020;83:104351.

